# Childhood growth trajectory patterns are associated with the pubertal gut microbiota

**DOI:** 10.1101/2023.06.20.23291663

**Authors:** Lorena López-Domínguez, Celine Bourdon, Jill Hamilton, Amel Taibi, Diego G Bassani, Juliana dos Santos Vaz, Romina Buffarini, Luciana Tovo Rodrigues, Iná S Santos, Alicia Matijasevich, M Carolina Borges, Anthony J Hanley, Aluísio JD Barros, Robert HJ Bandsma, Elena M Comelli

## Abstract

The relationship between growth and gut microbiota establishment is critical but under-investigated during late childhood. This is an important knowledge gap since the adult microbiota connects with other organs to influence health. We studied gut microbial communities composition and dietary patterns in 13 years old males and females from the 2004 Pelotas birth cohort (Brazil). We had previously shown that three BMIZ and HAZ patterns of growth since birth are present in this cohort, reflecting the nutritional status of these children. Here, we show that there is an association between growth patterns and gut microbiota, which encompasses the five pubertal stages, and which is affected by sex. Using Partial Least Square Path Modelling, we also show that there is a strong relationship between dietary patterns and gut microbiota, in males but not females. These data provide the microbiota and dietary profiles of highly phenotyped children and highlight the importance of childhood growth and sex for the maturation of the gut microbiota and long-term healthy growth. The role of pubertal diet in the relationships identified, further underlies the importance of dietary patterns establishment during adolescence while providing an opportunity for late modification of growth-microbiota relationships.

## INTRODUCTION

Children patterns of growth and health throughout life are intertwined. The first 3 years of life are critical for childhood growth and development and correspond to major diversification and maturation of the gut microbiota (Bokulich et al., 2016; Stewart et al., 2018; Yassour et al., 2018; Yatsunenko et al., 2012). Microbial colonization in early life plays a critical role in development and disturbances may impact health outcomes later in life (Robertson et al., 2019). It has been shown that undernourished children have a less mature gut microbiota than healthy children (Balasubramaniam et al., 2021; Méndez-Salazar et al., 2018; Subramanian et al., 2014). Studies in adequately nourished and over nourished children are scant. One study in Canadian children showed that preschool (0-5 years of age) BMI growth trajectories are associated with the longitudinal variation of the gut microbiota diversity and relative abundance at 3 and 12 months of age (Reyna et al., 2022).

Adolescence is the other critical period in childhood characterized by specific developmental milestones associated with rapid physical growth and sexual maturation (Norris et al., 2022). This is also an important time for dietary patterns modification (Winpenny et al., 2018) and dietary patterns are known to influence the gut microbiota (Singh et al., 2017; Turnbaugh et al., 2009; Wu et al., 2011). However, the relation between the gut microbiota, puberty and childhood growth has not been well studied. The microbiota in adolescence has been reported to differ from that of adults (Agans et al., 2011; Hollister et al., 2015; Ringel-Kulka et al., 2013; Yatsunenko et al., 2012). Children harbor a less diverse microbiota than adults (Agans et al., 2011; Radjabzadeh et al., 2020), and differences between sexes might be highlighted in puberty. Pubertal timing has been associated with microbiota composition in a sex-dependant manner and with hormone levels (Korpela et al., 2021; Yuan et al., 2020). No studies have explored the association between the gut microbiome and growth into adolescence.

The objective of this study was to assess whether childhood growth from birth is associated with the adolescent microbiota. We used a novel analytical pipeline that we recently established and that allows identification of growth trajectory patterns since birth while allowing for increased weight on later stages of childhood growth (López-Domínguez et al., 2023). We report that patterns of growth through childhood are associated with the gut microbiota during adolescence, suggesting that interventions to shift the trajectory to that of a healthy growth-compatible microbiome throughout early childhood could affect longer-term growth.

## METHODS

### Study design and participants

#### Pelotas 2004 Birth cohort

The 2004 Pelotas birth cohort is a prospective study of 4231 children born in 2004 in the urban area of Pelotas in southern Brazil (Barros et al., 2006). Briefly, mothers were recruited at the five maternity hospitals in the region, covering 98% of all deliveries, after providing written informed consent. After birth, children were followed up at 3, 12, 24, 48 months, and at 6 and 11 years of age when anthropometry, socioeconomic, behavioural, and demographic data were collected by trained research staff.

#### 12-year-old follow-up

In May 2017, the 2004 Pelotas Birth Cohort started a sub-study, the Oral Health Study, to assess the oral health of 1200 adolescents born between September and December of 2004 (Silveira Schuch et al., 2021). For our prospective cross-sectional study, we subsampled 500 children participating in the Oral Health Study (Figure 1). Children were stratified based on anthropometric z-scores calculated using data from the 11 years of age follow up with AnthroPlus Software by the World Health Organization (WHO 2009) and a stratified subsampling approach was used to ensure selection of participants from our subpopulations of interest (i.e., stunting, wasting, normal weight, overweight and obesity). All participants with height/age z-score below -2 (stunting) and those with BMI/age z-score below -2 (wasting) and above 3 (obesity) were included. The remaining participants were randomly selected in equal number from the following two strata: between -2 and 2 (normal weight) and between 2 and 3 (overweight) (Supplementary table 1).

**Figure 1.**
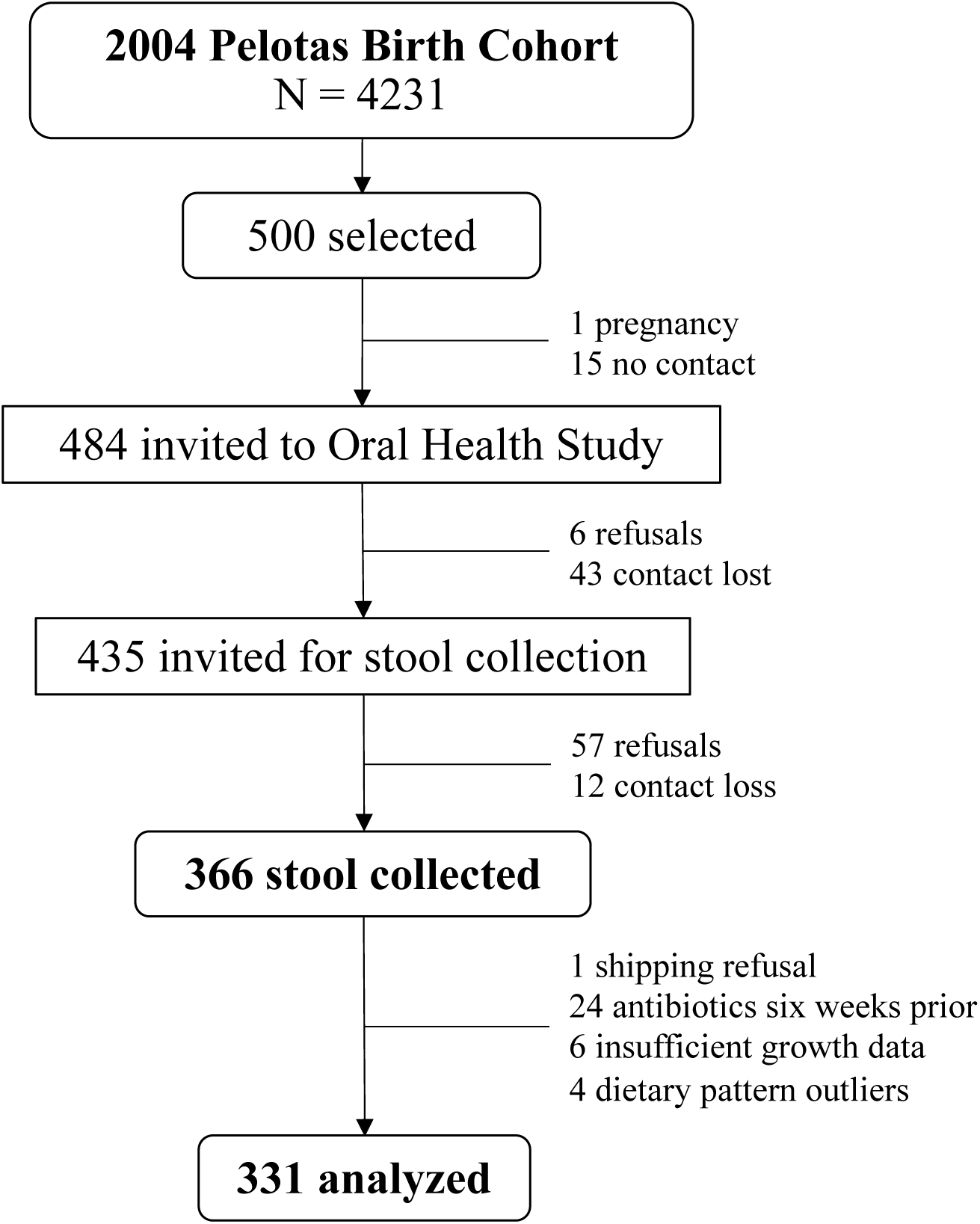
Flow diagram of sample selection and collection.

During their at-home visit, the Oral Health Study team invited children to participate, and those who agreed were provided with a stool collection kit. A follow-up home visit was carried out by a trained interviewer from the microbiome sub study to collect the stool sample, anthropometric measurements (weight, height, and abdominal circumference) and a questionnaire. The questionnaire collected information on pet ownership, physical activity and dietary habits, description of stool collection and handling, use of medications and included a pubertal-self assessment and a food frequency questionnaire (details are provided below).

### Anthropometric measurements and growth trajectories patterns

The details of anthropometric measurements were previously described (Barros et al., 2006; Santos et al., 2011, 2014). Briefly, children’s length/height and weight were measured using standardized protocols (Habicht, 1974) by trained personnel. Length/height at birth, 3, 12, 24 and 48 months was measured using a foldable wooden anthropometer (with 1 mm precision). Height at 6 and 11 years was taken with a stadiometer (Harpenden) (maximum 2.06 m and 1 mm precision). Birthweight was measured using electronic pediatric scales with 10 g precision, and subsequent measurements were taken using an electronic scale (150 kg capacity and 100 g precision). At 3, 12 and 24-month visits, mother’s and child’s weights were measured together, and the child’s final weight was calculated by subtracting mother’s weight and the estimated weight of any remaining clothes. At the 4, 6, 11 and 13-year visit, the child was weighted without shoes and wearing light clothes. Height and weight information collected at the 13 years follow up and used to derive Body Mass Index-for-age (BMIZ) and Height-for-age z-scores (HAZ), and then stratified based on WHO cut-offs. Growth trajectory patterns were derived from our previous study in this cohort, where we used shape-based clustering (k-means) of gestational-age corrected data, an approach that improves interpretability of growth patterns in later childhood (López-Domínguez et al., 2023).

### Pubertal assessment

Stages of puberty were identified according to Tanner (Marshall & Tanner, 1969, 1970) with a two-pages self-reported tool assessing breast and pubic hair development for females and genitalia and pubic hair development for males, developed based on previous literature (Morris et al. 1980) This method was previously shown to be in agreement with physician’s assessment (Matsudo & Matsudo, 1994; Rasmussen et al., 2015; Schmitz et al., 2004; Taylor et al., 2001). For subsequent analyses, children were grouped into three categories corresponding to Tanner Stages 1-2 (n=46), Stage 3 (n=104), and Stages 4-5 (n=184)).

### Dietary assessment

Children completed a semi-quantitative Food Frequency Questionnaire (FFQ) specifically developed for this population (Schneider et al., 2016; Vaz et al., 2021) that contains 87 food items classified into 11 groups and estimates food consumption during the previous year. The tool consists of a booklet with pictures of mean servings to estimate consumed portion sizes and frequency of consumption was recorded in a tablet. Eight frequency options were given for each food: (1) five or more times a day, (2) two to four times a day, (3) once a day, (4) five to six times a week, (5) two to four times a week, (6) once a week, (7) one to three times a month and (8) never or less than once a month. Energy and nutrient intake were analyzed using the Brazilian Table of Food Composition database (Lopes Tdo et al., 2015) complemented with the USDA Food Composition database (U.S. Department of Agriculture, 2017) for foods not included in the Brazilian database.

### Dietary patterns

Food items consumed by less than 20% of the participants were removed from the analysis, as they were considered not to be part of their regular diet (soy milk, nuts, shrimp, alcoholic drinks: beer, wine, other). The remaining items were combined into 19 groups based on nutrient composition, frequency of consumption and the population’s traditional diet (Supplementary table 2). To derive the dietary patterns, we applied Principal Components Analysis (PCA) and Varimax rotation to obtain orthogonal factors. The number of components extracted was based on eigenvalues >1.4 and scree test plots. Dietary patterns were named according to the food items included and participants received a score for each dietary pattern identified. Factor loadings of > 0.25 were considered to have strong associations with that pattern. Dietary pattern scores were classified into tertiles, and the group with the lowest score was considered to have the lowest adherence to that pattern.

### Stool sample collection

Stools were collected at home during the recruitment visit using the Stool Nucleic Acid Collection and Preservation Tube (Norgen Biotek^®^ Thorold, ON, Canada). A video was created to explain the collection process to the participants. The interviewer collected the samples at home and transported them to the Center of Epidemiology of the University of Pelotas for storage until shipping to the University of Toronto.

### Fecal microbiota composition analysis

DNA was extracted using the ZymoBIOMICS DNA Miniprep kit, product number D4300 (Zymo Research, Irvine, CA), according to the manufacturer’s instructions but including a lysis step (Mini BeadBeater (Biospecs Products Inc) run for 2 minutes). DNA was stored at -20°C. The V3V4 hypervariable region of the 16S rRNA gene was amplified from the DNA using the barcoded forward 338F (5’-ACTCCTACGGGAGGCAGCAG-3’) and reverse 806R (5’- GGACTACHVGGGTWTCTAAT-3’) sequencing primers to allow for multiplexing (Caporaso et al., 2012, Kozich et al., 2013). Amplification reactions were performed using 12.5 uL of KAPA2G Robust HotStart ReadyMix (KAPA Biosystems), 1.5 uL of 10 uM forward and reverse primers, 7.5 uL of sterile water and 2 uL of DNA. The V3V4 region was amplified by cycling the reaction at 95°C for 3 minutes, 18x cycles of 95°C for 15 seconds, 55°C for 15 seconds and 72°C for 15 seconds, followed by a 5-minute 72°C extension. All amplification reactions were done in triplicate to reduce amplification bias, pooled, and checked on a 1% agarose TBE gel. Pooled triplicates were quantified using PicoGreen and combined by even concentrations. The library was then purified using Ampure XP beads and loaded on to the Illumina MiSeq for sequencing, according to manufacturer instructions (Illumina, San Diego, CA). Sequencing is performed using the V3 (300bp x 2) chemistry. A single-species (*Pseudomonas aeruginosa* DNA), a mock community (Zymo Microbial Standard: https://www.zymoresearch.de/zymobiomics-community-standard) and a template-free negative control were included in the sequencing run.

The UNOISE pipeline, available through USEARCH v11.0.667 and vsearch v2.10.4, was used for sequence analysis (Edgar, 2010, 2013, 2016; Rognes et al., 2016). The last base was removed from all sequences, and the 3’ tail was trimmed after the quality dropped below q15 for each read, using cutadapt v.1.18. Sequences were assembled and quality trimmed using – fastq_mergepairs with a –fastq_trunctail set at 3, a –fastq_minqual set at 2, a -fastq_maxdiffs set at 20, a -fastq_pctid set at 70, and minimum and maximum assemble lengths set at 100 and 600 base pairs, respectively. Assembled sequences were quality filtered using –fastq_filter with a – fastq_maxee set at 1.0. The trimmed data was then processed following the UNOISE pipeline. Sequences were first de-replicated and sorted to remove singletons, then denoised and chimeras were removed using the unoise3 command. Assembled sequences were mapped back to the chimera-free denoised sequences at 99% identity OTUs. Taxonomy assignment was executed using SINTAX, available through USEARCH, and the UNOISE compatible Ribosomal Database Project (RDP) database version 16, with a minimum confidence cutoff of 0.8 (Wang et al., 2007). OTU sequences were aligned using align_seqs.py v.1.9.1 through QIIME1 (Caporaso et al., 2010). Sequences that did not align were removed from the dataset and a phylogenetic tree of the filtered aligned sequence data was made using FastTree (Price et al., 2009). The 16S copy number were estimated with the SINAPS algorithm, accessed through USEARCH. These analyses were performed at the University of Toronto Centre for the Analysis of Genome Evolution & Function (CAGEF). Mean sequencing depth of samples was 29,742 (2,120 - 67,121) and data were rarefied at 2,000 sequences per sample to calculate diversity metrics. Alpha-diversity (Chao1 and Shannon index (Chao, 1984; Shannon, 1948)) and beta-diversity metrics (weighted and unweighted UniFrac distances (C. A. Lozupone et al., 2007; C. Lozupone & Knight, 2005)) were estimated in QIIME2 using q2-diversity (Bolyen et al., 2019).

### Statistical analyses

Associations between the gut microbiota diversity and variables of interest were assessed as follows: Within groups diversity (alpha diversity) was assessed using Kruskal-Wallis and generalized linear models with microbiota richness (Chao1 index) and evenness (Shannon index). Variance between groups (beta diversity) was assessed with Permutation analysis of variance (PERMANOVA) and multivariate ANOVA based on similarity tests (ADONIS) (Anderson, 2001) using Unweighted and Weighted UNIFRAC distance metrics. Models were constructed by adjusting for sex, skin colour, household income, type of birth, and perinatal characteristics (maternal age, education and smoking during pregnancy) selected after testing them independently for associations with the microbiota (Supplementary figure 1 and supplementary tables 3 and 4). Differential abundance was assessed with analysis of compositions of microbiomes (ANCOM) (Mandal et al., 2015) with OTUs.

### Partial Least Squares Path Modeling (PLS-PM)

PLS-PM was used to identify the relationships between growth trajectories since birth, pubertal status, dietary patterns, and gut microbiota composition using the plspm R package (Sanchez, 2013). PLS is an exploratory method that allows to test complex relationships among variables using latent variables and uses bootstrapping to obtain information about the variability of the parameter estimates. We created a model of cross correlations integrating microbiota profiles and the explanatory variables and identify the key OTUs that were responsible for the differential microbiota structure. Models were tested for the whole sample and separately by growth pattern and by sex.

## RESULTS

### Participant characteristics and growth patterns membership

331 children, 12.8 ± 0.17 years old, were included in the study (Figure 1). Participants were classified into wasting (n=11), normal (n=245) and overweight (n=75) for BMIZ; and in stunting (n=10) and normal (n=321) for HAZ (Table 1). Females (46.5%) were predominantly classified in pubertal stage 4-5 (70.8%), followed by stage 3 (19.5%) and stages 1-2 (9.7%). In contrast, males were equally classified into stage 3 (41.8%) and stages 4-5 (40.1%), followed by stages 1-2 (18.1%). Most children were grouped into the Stable growth trajectories pattern (BMIz n = 157, 47%; HAz n = 165, 50%), followed by the Decreasing (BMIz n = 96, 29%; HAz n = 98, 30%), and the Increasing (BMIz n = 78, 24%; HAz n = 68, 20%) patterns. Mean BMIz and HAz were classified as “normal” in all patterns, except for the BMIz Increasing pattern. The distribution among patterns of children selected for this study reflects that of the whole cohort for both the BMIz and HAz (López-Domínguez et al., 2023).

**Table 1.** Participants characteristics by growth trajectory pattern. The growth trajectory patterns are as per our previous study (López-Domínguez et al., 2023).

|  | ALL<br>(n=331) | BMIz pattern |  |  | HAz pattern |  |  |
| --- | --- | --- | --- | --- | --- | --- | --- |
|  |  | Decreasing<br>(n=96) | Stable<br>(n=157) | Increasing<br>(n=78) | Decreasing<br>(n=98) | Stable<br>(n=165) | Increasing<br>(n=68) |
| **Age (years)** | 12.8 ± 0.17 | 12.8 ± 0.16 | 12.8 ± 0.17 | 12.8 ± 0.18 | 12.8 ± 0.18 | 12.8 ± 0.17 | 12.8 ± 0.15 |
| **Sex (Female)** | 154 (46.5%) | 50 (52.1%) | 73 (46.5%) | 31 (39.7%) | 44 (44.9%) | 78 (47.3%) | 32 (47.1%) |
| **Maternal age (years)** | 25.9 ± 6.85 | 25.9 ± 6.94 | 25.7 ± 6.92 | 26.2 ± 6.64 | 25.7 ± 6.73 | 26.1 ± 7.1 | 25.5 ± 6.43 |
| **Maternal education (years)** | 8.2 ± 3.25 | 7.5 ± 3.15 | 8 ± 3.18 | 9.2 ± 3.29 | 7.2 ± 3.1 | 8.5 ± 3.18 | 8.7 ± 3.38 |
| **Maternal BMI (kg/m2)** | 23.8 ± 4.53 | 23.4 ± 4.34 | 23.5 ± 4.71 | 24.6 ± 4.39 | 24.3 ± 5.3 | 23.4 ± 4.19 | 24 ± 4.16 |
| **Maternal smoking (yes)** | 84 (25.4%) | 22 (22.9%) | 39 (24.8%) | 23 (29.5%) | 32 (32.7%) | 41 (24.8%) | 11 (16.2%) |
| **Skin colour** |  |  |  |  |  |  |  |
| *White* | 210 (63.4%) | 60 (62.5%) | 95 (60.5%) | 55 (70.5%) | 58 (59.2%) | 107 (64.8%) | 45 (66.2%) |
| *Black* | 52 (15.7%) | 19 (19.8%) | 23 (14.6%) | 10 (12.8%) | 16 (16.3%) | 27 (16.4%) | 9 (13.2%) |
| *Brown* | 53 (16%) | 11 (11.5%) | 30 (19.1%) | 12 (15.4%) | 16 (16.3%) | 26 (15.8%) | 11 (16.2%) |
| *Other* | 12 (3.6%) | 3 (3.1%) | 8 (5.1%) | 1 (1.3%) | 7 (7.1%) | 3 (1.8%) | 2 (2.9%) |
| **Income** |  |  |  |  |  |  |  |
| *Quintile 1* | 64 (19.3%) | 18 (18.8%) | 30 (19.1%) | 16 (20.5%) | 18 (18.4%) | 32 (19.4%) | 14 (20.6%) |
| *Quintile 2* | 72 (21.8%) | 21 (21.9%) | 38 (24.2%) | 13 (16.7%) | 20 (20.4%) | 38 (23%) | 14 (20.6%) |
| *Quintile 3* | 55 (16.6%) | 11 (11.5%) | 26 (16.6%) | 18 (23.1%) | 22 (22.4%) | 24 (14.5%) | 9 (13.2%) |
| *Quintile 4* | 81 (24.5%) | 29 (30.2%) | 33 (21%) | 19 (24.4%) | 25 (25.5%) | 41 (24.8%) | 15 (22.1%) |
| *Quintile 5* | 59 (17.8%) | 17 (17.7%) | 30 (19.1%) | 12 (15.4%) | 13 (13.3%) | 30 (18.2%) | 16 (23.5%) |
| **Type of birth (cesarian)** | 154 (46.5%) | 41 (42.7%) | 71 (45.2%) | 42 (53.8%) | 37 (37.8%) | 74 (44.8%) | 43 (63.2%) |
| **Breastfeeding at 3 months** |  |  |  |  |  |  |  |
| *No* | 73 (22.1%) | 21 (21.9%) | 39 (24.8%) | 13 (16.7%) | 24 (24.5%) | 32 (19.4%) | 17 (25%) |
| *Partial* | 111 (33.5%) | 30 (31.2%) | 50 (31.8%) | 31 (39.7%) | 33 (33.7%) | 61 (37%) | 17 (25%) |
| *Predominant* | 54 (16.3%) | 19 (19.8%) | 27 (17.2%) | 8 (10.3%) | 16 (16.3%) | 26 (15.8%) | 12 (17.6%) |
| *Exclusive* | 93 (28.1%) | 26 (27.1%) | 41 (26.1%) | 26 (33.3%) | 25 (25.5%) | 46 (27.9%) | 22 (32.4%) |
| **Breastfeeding at 12 months (yes)** | 139 (42%) | 38 (39.6%) | 69 (43.9%) | 32 (41%) | 39 (39.8%) | 72 (43.6%) | 28 (41.2%) |
| **Age of introduction of solids (months)** | 4.4 ± 1.28 | 4.5 ± 1.33 | 4.4 ± 1.28 | 4.5 ± 1.23 | 4.4 ± 1.3 | 4.4 ± 1.27 | 4.4 ± 1.29 |
| **BMI-for-age z-score** | 0.7 ± 1.56 | -0.3 ± 1.3 | 0.6 ± 1.43 | 2 ± 1.19 | 0.5 ± 1.42 | 0.5 ± 1.65 | 1.1 ± 1.45 |
| *Wasting (<-2 BMIz)* | 11 (3.3%) | 6 (6.2%) | 5 (3.2%) | 0 (0%) | 0 (0%) | 11 (6.7%) | 0 (0%) |
| *Overweight (>2 BMIz)* | 75 (22.7%) | 3 (3.1%) | 28 (17.8%) | 44 (56.4%) | 18 (18.4%) | 33 (20%) | 24 (35.3%) |
| **Height-for-age z-score** | 0.3 ± 1.14 | 0 ± 1.07 | 0.3 ± 1.14 | 0.7 ± 1.11 | -0.2 ± 1.06 | 0.3 ± 1.05 | 1 ± 1.14 |
| *Stunting (<-2 HAz)* | 10 (3%) | 5 (5.2%) | 4 (2.5%) | 1 (1.3%) | 6 (6.1%) | 4 (2.4%) | 0 (0%) |
| **Fat mass at 11 years (%)** | 28.6 ± 12.56 | 21.1 ± 9.03 | 28.3 ± 11.92 | 38.3 ± 11.1 | 26.7 ± 11.69 | 28.2 ± 13.01 | 32.2 ± 12.1 |
| **Pubertal stage Females** |  |  |  |  |  |  |  |
| *Stage 1-2* | 15 (9.7%) | 10 (20%) | 4 (5.5%) | 1 (3.2%) | 3 (6.8%) | 9 (11.5%) | 3 (9.4%) |
| *Stage 3* | 30 (19.5%) | 12 (24%) | 13 (17.8%) | 5 (16.1%) | 8 (18.2%) | 19 (24.4%) | 3 (9.4%) |
| *Stage 4-5* | 109 (70.8%) | 28 (56%) | 56 (76.7%) | 25 (80.6%) | 33 (75%) | 50 (64.1%) | 26 (81.2%) |
| **Pubertal stage Males** |  |  |  |  |  |  |  |
| *Stage 1-2* | 32 (18.1%) | 6 (13%) | 17 (20.2%) | 9 (19.1%) | 15 (27.8%) | 10 (11.5%) | 7 (19.4%) |
| *Stage 3* | 74 (41.8%) | 21 (45.7%) | 33 (39.3%) | 20 (42.6%) | 23 (42.6%) | 32 (36.8%) | 19 (52.8%) |
| *Stage 4-5* | 71 (40.1%) | 19 (41.3%) | 34 (40.5%) | 18 (38.3%) | 16 (29.6%) | 45 (51.7%) | 10 (27.8%) |

### Dietary patterns

Three dietary patterns were identified to best describe the reported dietary intake of participants, explaining a cumulative 35% of the total variance (Table 2). The first component identified was labelled “Processed”, and contained high loadings of fast foods and snacks, candies and sweets, processed meats, cookies and cake, pastas and tubers, soda, and dairy. The second component labelled “Coffee, Tea and Vegetables” was characterized by high loadings of coffee, tea, sugar, and vegetables and legumes. The third component was labelled “common-Brazilian”, with high loadings of beans, rice, bread, and fats. The percentage of variance explained by each dietary pattern was 13.1%, 11.6%, and 10.8%, respectively. The eigenvalues in each dietary pattern were 3.4 (Processed), 1.8 (Coffee, Tea and Vegetables), and 1.4 (common-Brazilian).

**Table 2.** Factor loadings of the three dietary patterns identified. The dietary patterns were composed by the food groups with the value of the factor loadings in bold.

|  | **Processed** | **Coffee, Tea and Vegetables** | **Common-Brazilian** |
| --- | --- | --- | --- |
| Fast food and snacks | **0.446** | 0.062 | -0.142 |
| Candies and sweets | **0.410** | -0.170 | 0.191 |
| Milk and dairy products | **0.356** | -0.304 | 0.099 |
| Processed meats | **0.335** | 0.037 | 0.047 |
| Cookies and cake | **0.320** | 0.088 | -0.146 |
| Pastas and tubers | **0.287** | 0.204 | -0.200 |
| Soda | **0.251** | -0.063 | -0.187 |
| Coffee | -0.033 | **0.557** | 0.031 |
| Sugar | 0.042 | **0.499** | 0.037 |
| Teas | -0.072 | **0.337** | -0.135 |
| Vegetables and legumes | 0.086 | **0.307** | 0.065 |
| Beans | -0.052 | -0.014 | **0.492** |
| Rice | -0.006 | -0.017 | **0.456** |
| Bread | 0.027 | 0.078 | **0.381** |
| Fats | 0.099 | 0.077 | **0.323** |
| Meats and eggs | 0.218 | 0.095 | 0.119 |
| Fruits and fruit juices | 0.196 | 0.125 | 0.202 |
| Fish | 0.176 | 0.048 | -0.046 |
| Vegetable spices | -0.012 | 0.116 | 0.232 |
| Number of food groups | 7 | 4 | 4 |
| Eigenvalues | 3.43 | 1.84 | 1.48 |
| % of variance explained | 13.1% | 11.6% | 10.8% |
| % of cumulative variance explained | 13.1% | 24.7% | 35.5% |

### Gut microbiota

Fourteen unique phyla-level and 157 unique genus-level taxa were identified (Figure 2). Based on relative abundance, at the phylum level the gut microbiota consisted primarily of Firmicutes (62.3 ± 16%) and Bacteroidetes (30.1 ± 17%), followed by Actinobacteria and Proteobacteria (3.9 ± 4% and 1.5 ± 2%, respectively); only these top four phyla were present in all samples (Figure 2A). There was a gradient, and an inverse relationship, in the relative abundance of Firmicutes and Bacteroidetes, with their ratio varying between 0 - 53.7 and 0 - 34.4 across the sample. Sixty percent of all stool samples (n=200) contained Archaea.

**Figure 2.**
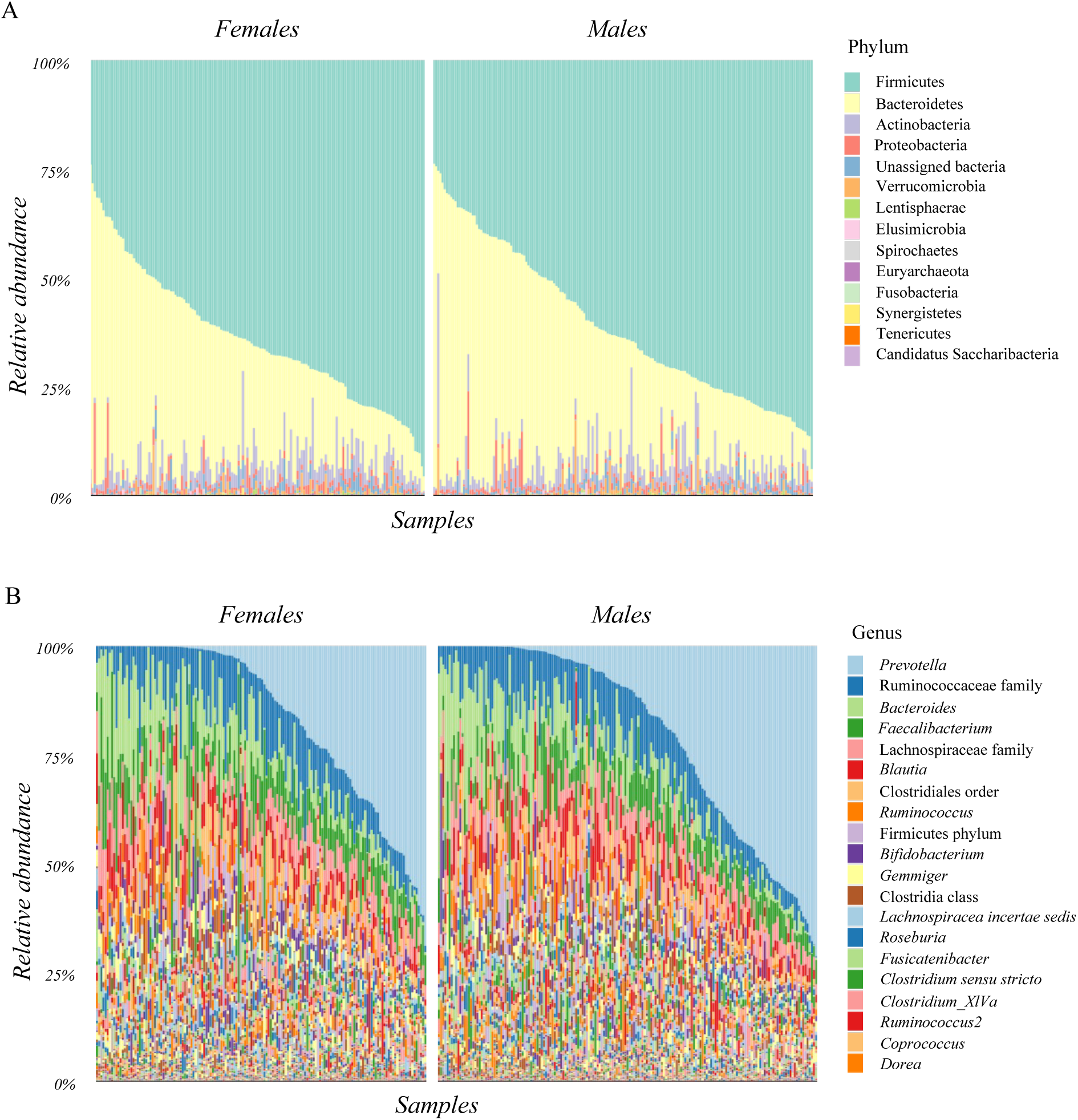
Gut microbiota from a subsample of children from the 2004 Pelotas Birth Cohort at 13 years of age. Relative abundance at the A) phylum and B) genus level (top 20 genera shown) by sex.

At the genus level, *Prevotella* (18.1 ± 20%), unidentified-genus from the Ruminococcaceae family (8.9 ± 6%), *Bacteroides* (8.5 ± 9%), and *Faecalibacterium* (8 ± 9%) were predominant, followed by unidentified genus from the *Lachnospiraceae* family (6.2 ± 4%), *Blautia* (5 ± 4%), unidentified genus from the Clostridiales order (4.6 ± 3%), and *Ruminococcus* (3.1 ± 3%) (Figure 2B). Only 7 genera were found in all samples: unidentified genus from the Ruminococcaceae family, *Bacteroides*, *Faecalibacterium*, unidentified genus from the *Lachnospiraceae* family, *Blautia*, unidentified genus from the Clostridiales order, and *Lachnospiracea incertae sedis* (1.8 ± 1%).

### Gut microbiota diversity, sex, growth patterns since birth, pubertal stage and diet

#### Sex

No differences in gut microbial alpha or beta diversity were observed among sex in the crude or adjusted models (Figure 3A and Table 3).

**Figure 3.**
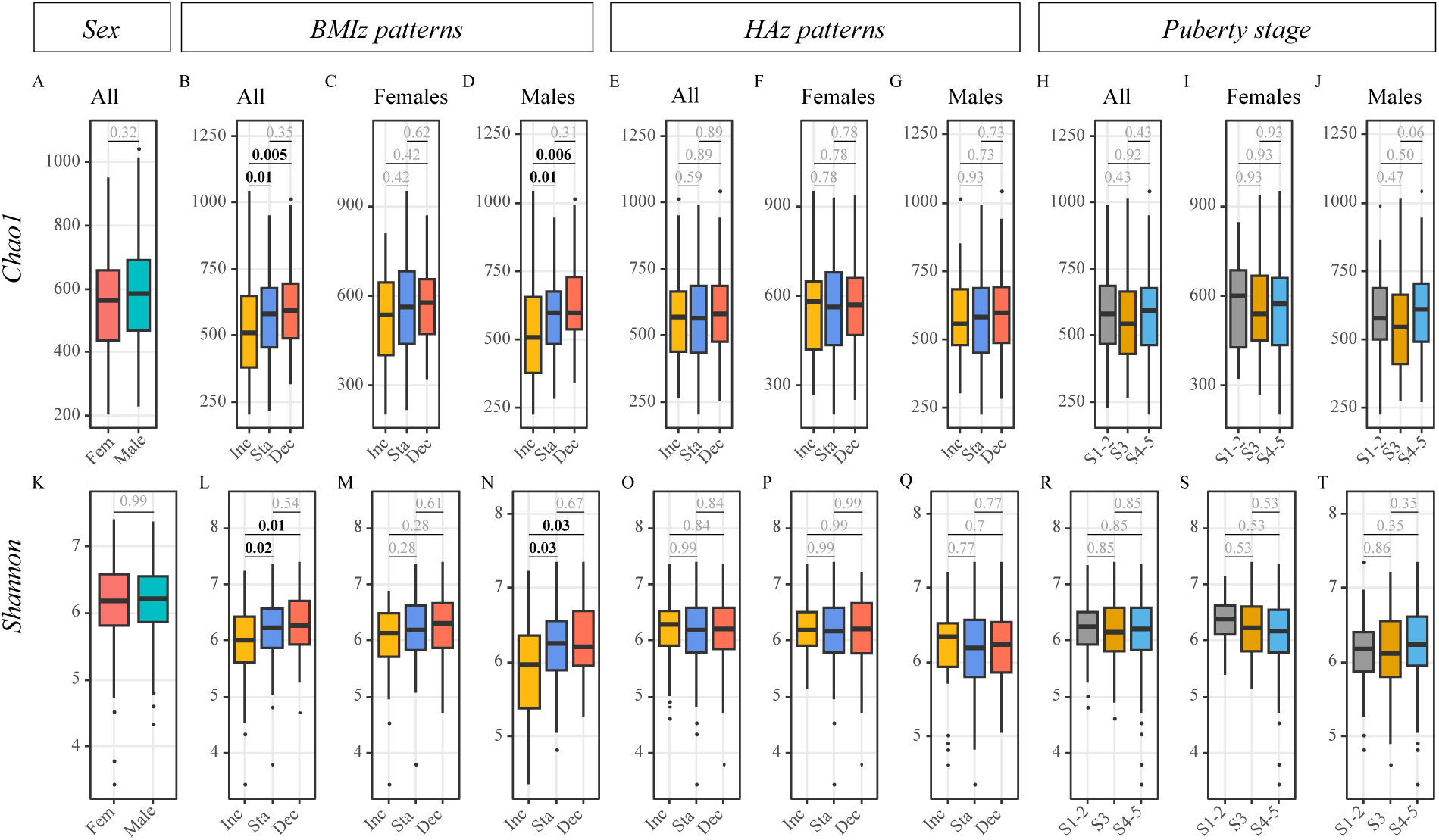
Association of sex, puberty stage, BMIz and HAz patterns, and the gut microbiota alpha diversity. Gut microbiota bacterial richness (Chao1 index) and evenness (Shannon index) in association with Sex (A, K), BMIz patterns (B, C, D, L, M, N), HAz patterns (E, F, G, O, P, Q), and Puberty stage (H, I, J, R, S, T). Associations were tested using Kruskal-Wallis test with significance at p < 0.05 (bold). Abbreviations: NS, not significant; Fem, female; Inc, increasing; Sta, stable; Dec, decreasing.

**Table 3.** Comparison (p-values shown) of gut microbiota beta-diversity and variables of interest (sex, puberty, BMIz and HAz patterns, dietary patterns). Associations were tested using PERMANOVA test (999 permutations) where significance was set at p < 0.05.

|  | Unweighted UNIFRAC |  |  | Weighted UNIFRAC |  |  |
| --- | --- | --- | --- | --- | --- | --- |
|  | All<br>(n=331) | Females<br>(n=154) | Males<br>(n=77) | All<br>(n=331) | Females<br>(n=154) | Males<br>(n=77) |
| **Sex (Female)** | 0.67 |  |  | 0.08 |  |  |
| **Puberty stage** | 0.757 | 0.739 | 0.057 | 0.646 | 0.876 | 0.591 |
| *Stage 1-2 vs 3* | 0.86 | 0.9 | 0.43 | 0.87 | 0.79 | 0.84 |
| *Stage 1-2 vs 4-5* | 0.86 | 0.9 | 0.64 | 0.87 | 0.79 | 0.84 |
| *Stage 3 vs 4-5* | 0.86 | 0.9 | **0.02** | 0.87 | 0.79 | 0.8 |
| **BMIz pattern** | **0.009** | 0.21 | 0.05 | 0.12 | 0.23 | 0.71 |
| *Decreasing vs Stable* | 0.89 | 0.94 | 0.86 | 0.45 | 0.9 | 0.79 |
| *Decreasing vs Increasing* | **0.001** | 0.12 | 0.05 | 0.13 | 0.19 | 0.79 |
| *Stable vs Increasing* | **0.001** | 0.16 | 0.05 | 0.21 | 0.24 | 0.79 |
| **HAz pattern** | 0.08 | 0.06 | 0.34 | 0.2 | 0.38 | 0.45 |
| *Decreasing vs Stable* | 0.24 | 0.17 | 0.7 | 0.32 | 0.62 | 0.46 |
| *Decreasing vs Increasing* | 0.09 | **0.02** | 0.44 | 0.23 | 0.56 | 0.46 |
| *Stable vs Increasing* | 0.45 | 0.58 | 0.37 | 0.49 | 0.66 | 0.46 |
| **Dietary pattern adherence** |  |  |  |  |  |  |
| **Processed** | 0.4 | 0.34 | 0.34 | 0.8 | 0.35 | 0.32 |
| *High vs Low* | 0.39 | 0.44 | 0.37 | 0.78 | 0.41 | 0.43 |
| *High vs Medium* | 0.52 | 0.57 | 0.59 | 0.78 | 0.41 | 0.43 |
| *Low vs Medium* | 0.94 | 0.57 | 0.59 | 0.78 | 0.58 | 0.58 |
| **Coffee, Tea and Vegetables** | **0.001** | **0.003** | **0.003** | **0.009** | 0.19 | 0.18 |
| *High vs Low* | **0.003** | **0.009** | **0.01** | **0.02** | 0.14 | 0.13 |
| *High vs Medium* | **0.03** | **0.01** | **0.02** | 0.71 | 0.56 | 0.57 |
| *Low vs Medium* | **0.02** | 0.63 | 0.61 | **0.03** | 0.6 | 0.58 |
| **Common-Brazilian** | **0.001** | **0.001** | **0.001** | **0.006** | **0.03** | **0.03** |
| *High vs Low* | **0.003** | **0.003** | **0.003** | **0.01** | **0.02** | **0.03** |
| *High vs Medium* | **0.02** | 0.08 | 0.07 | 0.45 | 0.35 | 0.35 |
| *Low vs Medium* | 0.07 | 0.1 | 0.12 | **0.01** | 0.16 | 0.14 |

#### BMIZ patterns

Children in the Increasing BMIz pattern had lower richness (Chao1 Kruskal-Wallis, p< 0.001) and evenness (Shannon Kruskal-Wallis, p< 0.01) compared to those from the Stable and Decreasing patterns (Figure 3). The gut microbiota beta-diversity of children in the Increasing BMIz pattern was different from those in the Decreasing and Stable patterns only when microbial abundance was not accounted for (Unweighted UniFrac PERMANOVA, p<0.01) (Table 3). Subgroup analysis showed that differences were only observed in male participants. Differences remained in the adjusted model.

#### HAZ patterns

No differences in gut microbial alpha or beta diversity were observed among HAz trajectory patterns in the crude or adjusted models (Figure 3 E-G, 3 O-Q and Table 3).

#### Pubertal stage

No differences in gut microbial alpha or beta diversity were observed among pubertal stages in the crude or adjusted models (Figure 3 H-J, 3 R-T and Table 3).

#### Dietary patterns

Children with low adherence to the common-Brazilian dietary pattern displayed lower richness (Chao1 Kruskal-Wallis, p< 0.05) and evenness (Shannon Kruskal-Wallis, p< 0.05) compared to those with high adherence (Figure 4). Different gut microbiota was displayed compared to both medium and high adherence groups (Unweighted and Weighted UniFrac PERMANOVA, p<0.05). In the subgroup analysis, differences in richness and evenness were observed only in females. The gut microbiota beta-diversity was also different between all tertiles of adherence for the Coffee, Tea and Vegetables pattern when abundance was not accounted for (Unweighted UniFrac PERMANOVA, p<0.01) (Table 3) and lower richness was found in the low adherence group compared to the high adherence (Figure 4). In the subgroup analysis, differences were shown in males only between the High and the Middle and Low tertiles. No differences in evenness were observed in this pattern. We were not able to identify any differences in the gut microbiota alpha (Figure 4 A-C, 4 J-L) or beta diversity (Table 3), when looking at the Processed dietary pattern. Differences remained the same in the adjusted models.

**Figure 4.**
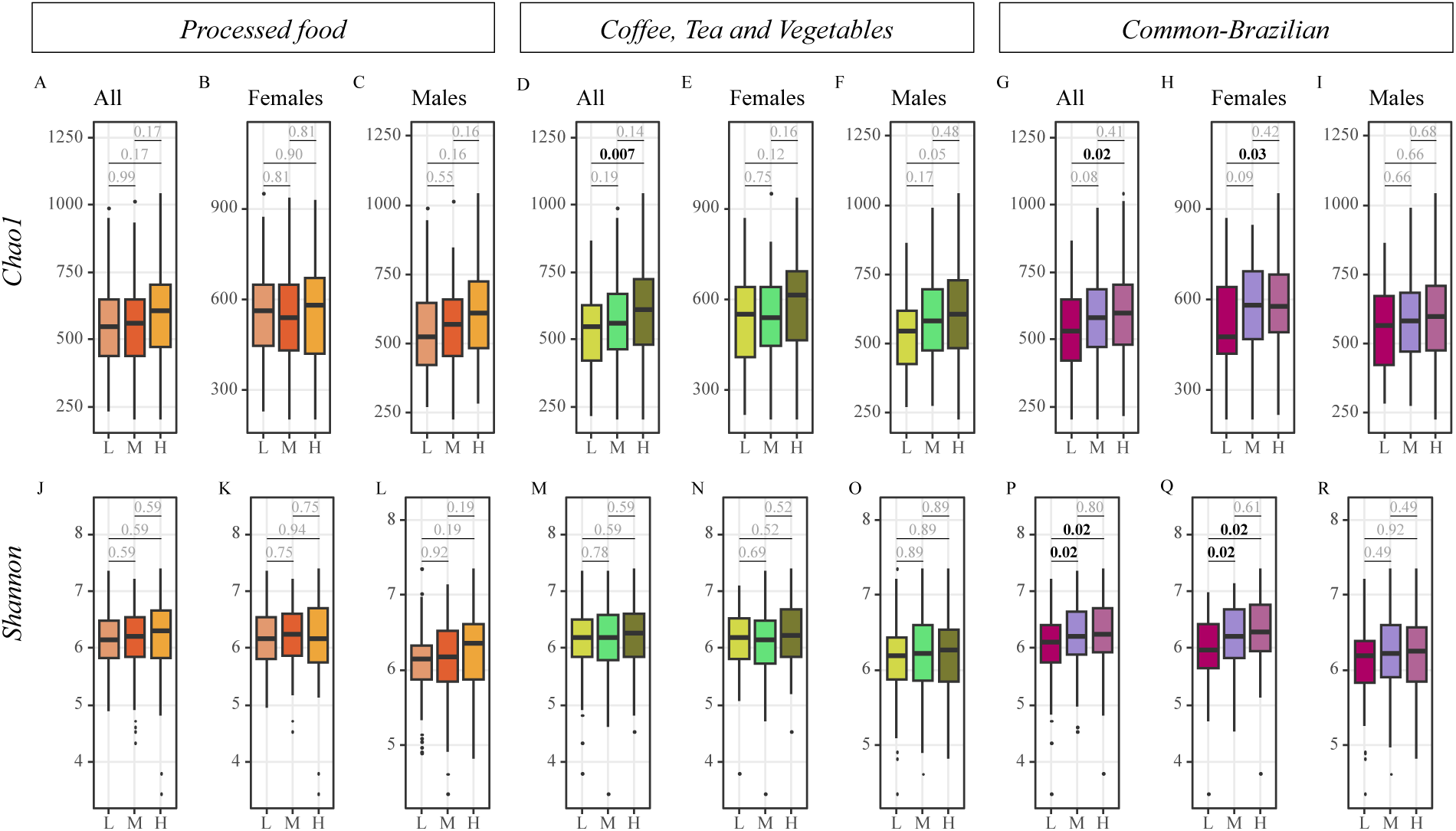
Association of dietary patterns and the gut microbiota alpha diversity. Gut microbiota bacterial richness (Chao1 index) and evenness (Shannon index) in association with food patterns adherence: Processed Food, A, B, C, J, K, L ; Coffee, Tea and Vegetables, E, F, G, O, P, Q; and Common-Brazilian, H, I, J, R, S, T. Associations were tested using Kruskal-Wallis test with significance at p < 0.05 (bold). Abbreviations: L, Lower tertile; M, Mid tertile, H, Higher tertile.

### Gut microbiota composition, sex, growth patterns since birth, pubertal stage and diet

#### Sex

We did not identify significant features among sex.

#### BMIZ patterns

We did not identify significant features among BMIz patterns.

#### HAZ patterns

We did not identify significant features among HAz patterns.

#### Pubertal stage

We did not identify significant features among pubertal stages.

#### Dietary patterns

For the Processed dietary pattern females *Prevotella* was higher in those with low adherence. Children with high adherence to the Coffee, Tea and Vegetables pattern had lower abundance of *Clostridium XIVa*, Ruminococcaceae family, *Prevotella*, and higher of *Prevotella copri*. In those with low adherence, Firmicutes was lower, and those with medium adherence had higher abundance of Clostridiales order. Specifically, in females *Clostridium XIVa* was lower in the high adherence group, and in males Firmicutes was lower in the low adherence group. Low adherence to the common-Brazilian pattern was associated with lower *Lactobacillus ruminis* in the whole sample, and with *Faecalibacterium prausnitzii* in females.

### Path modeling

To analyze the relationship among microbiota composition and the growth patterns, puberty, and dietary patterns, PLS-PM were constructed for the whole sample and separately by sex for the BMIz and HAz patterns (Figure 5). We hypothesized that the association between growth trajectory patterns in childhood and pubertal stage is at least partly explained by the correlation between dietary patterns and the gut microbiota composition. In the BMIz model, negative associations were shown between dietary patterns and BMIz patterns with the gut microbiota in the sex-specific models (Females -0.22, -0.22; Males -0.50, -0.22), in this model the gut microbiota was negatively associated with puberty (-0.22). This relationship was also observed in the by-sex analysis, though the negative association was found only in females (-0.34, Figure 5C), while a positive association was found in the males model (0.16, Figure 5E). In all paths modeled the dietary patterns, BMIz and HAz growth patterns showed an association with the microbiota composition, and in turn the microbiota composition influencing puberty stage, although the direction of the relationship was different between sex (Supplementary tables 5 and 6).

**Figure 5.**
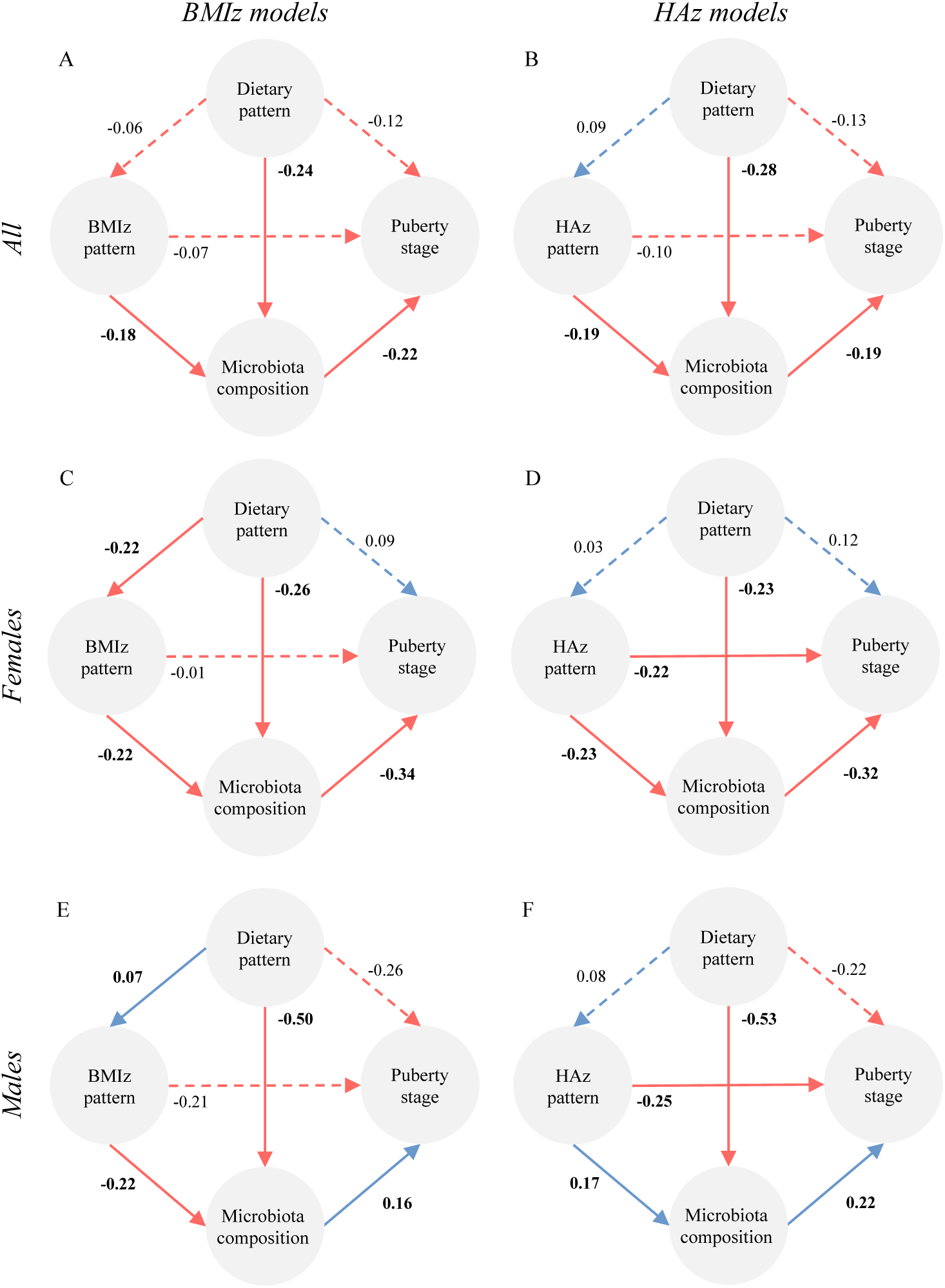
Cross-associations between childhood growth patterns, dietary patterns, pubertal stage and microbiota composition at 13 years of age. Arrows represent the associated paths tested between the BMIz and HAz growth patterns, dietary patterns loadings, pubertal stages and microbiota composition summarized as OTUs for A, B) the whole sample, C, D) Females and E, F) males. Numbers indicate the path coefficients representing the strength and direction of the relations between the response and the predictors. Blue and red lines represent positive and negative effects, respectively. Dashed lines indicate the coefficients that did not significantly (P > 0.05) differ from 0.

## DISCUSSION

The host and its gut microbiota co-evolve in early life to reach a mature dynamic equilibrium status that characterizes adulthood. Although it is recognized that very early life, within 3 years of age, is a critical stage for the formation of the microbiota, research in recent years suggests that its continuous reshaping during childhood growth may be critical to establish the adult baseline. Here we show that there is indeed an association between growth trajectory patterns from birth to late childhood and the gut microbiota at age 13. We also show that the relationship encompasses stages of puberty and dietary patterns, and that it is, at least in part, sex-specific. Although the sex variable was not associated with the gut microbiota in this study, even in the adjusted models, differences observed among variables were sex dependant.

Sex specificity of gut microbial communities is an ongoing topic of research and data suggests that sex is an important contributor in shaping the gut microbiota in early life (Kim et al., 2020; Valeri & Endres, 2021). Pre-clinical studies were the first to provide evidence that the microbiota dichotomizes by sex at puberty (Markle et al., 2013). The concept has been more recently investigated in two human studies, one in 5-15 years old Chinese children (Yuan et al., 2020) and one in 13 years old Finnish children (Korpela et al., 2021), both using 16S rRNA sequencing data. The first found taxa associated with levels of testosterone and the second found that pubertal timing was associated with the microbiota in females but not in males. We did not find any differences in terms of alpha and beta diversity, nor taxa relative composition between males and females. Because the microbiota and hormone metabolism, including estradiol and testosterone, are related (Calcaterra et al., 2022; Shin et al., 2019), assessing hormonal levels in the children under study may be necessary to reconcile results across studies. Towards the understanding of potential hormone-dependent relationships, we investigated whether the microbiota differed among pubertal stages but were not able to detect differences. Similar to Yuan et al, this could be due to the age of the children in our study (most females (76%) had already reached menarche), and to its cross-sectional nature. Although, as expected (WHO, 2010), there was a larger spread across pubertal stages in boys, the sample size within the subgroups, as well as inter-individual variation, was likely too low to detect potential differences.

We found that children, specifically males, with an Increasing BMIz trajectory pattern have a distinct microbiota diversity, characterized by lower evenness and richness, compared to those who grew in a Stable or Decreasing pattern. Similar results were reported in a Canadian cohort (Reyna et al., 2022), where children grouped in a rapid BMIz growth trajectory gained less microbiota diversity in their first year of life. A cohort study in Norway reported that the gut microbiota at age 2 has the potential to identify children at risk of obesity later in life (Stanislawski et al., 2018). In the present study, the Increasing BMIz pattern also showed higher proportion of children classified as overweight at 13 years and higher fat mass at 11 years than those in the other two patterns. Lower diversity has been found in individuals with obesity (Castaner et al., 2018) and a recent review highlighted specific taxa associated with obesity in school-age children (Vander Wyst et al., 2021). We were not able to identify specific taxa associated with the growth trajectories but found lower richness in children classified as overweight at 13 years, and that this difference was driven by males.

Most studies carried out in adolescents do not include the effect of dietary intake. This is important because adolescence comprises a stage of dietary independence and because of the heavy influence diet has on shaping the gut microbiota (Singh et al., 2017; Turnbaugh et al., 2009; Wu et al., 2011). Here, we showed that children, specifically females, with high adherence to the common-Brazilian dietary pattern, characterized for high intake of low processed foods, have higher microbiota evenness and richness. Although we did not find associations with the Processed dietary pattern, the microbiota from children with high adherence to both the common-Brazilian and the Vegetable, Tea and Coffee patterns differed from those with low adherence. Also, the path analyses showed the strongest relation to be between the dietary pattern and the microbiome in males, specifically.

This is the first study to assess the association of childhood growth trajectories and pubertal microbiota. We were able to integrate fecal sample collection in an established birth cohort and include perinatal and childhood variables in the analysis. Thanks to the great follow up rates from the Pelotas Birth cohorts, we were able to collect a large number of samples. Some limitations include the assessment of pubertal staging with a self-reported tool, which could lead to individual bias and is less reliable measuring hormonal levels in blood. The use of a FFQ relies on child’s memory and thus is prone to bias, we tried to overcome this by using PCA instead of only focusing on specific nutrients. Due to the high proportion of girls who had reached puberty in this study, the age of follow-up might not be ideal to study pubertal variations, new research should try to include a wide age range of participants to better capture the different stages.

We identified associations among childhood growth, diet the pubertal gut microbiota in a sex-dependent manner. This means that we could intervene throughout early childhood to affect the microbiome in puberty, and beyond this time, we may still have an opportunity to shift the trajectory to that of a healthy compatible microbiome. While microbiota is known to respond to short-term stimuli, core microbial features seem to be established through years and the entire period of childhood is important. Further research should focus on clarifying the direction of the associations among gut microbiota, growth, puberty and diet.

## Data Availability

All data that support the findings of this study are available from the 2004 Pelotas Birth cohort study investigators, but restrictions apply to the availability of these data, which were used under license for the current study, and so are not publicly available. Data are however available upon reasonable request and with permission of the 2004 Pelotas Birth cohort research committee.
Analytical code will be shared upon publication in a peer reviewed journal.

## ACKNOWLEDGEMENTS

The authors thank all the participants, researchers and the team involved in the 2004 Pelotas Birth Cohort. We also thank Marina Carpena, Thais Martins, Nathalia Victoria Pinto da Silva, Mariana Xavier, Rafaela Martins and Francine Costa for their participation in the sub study fieldwork. We also thank Drs. Pauline Wang and Sylva Donaldson for sample management and processing, and Dr. Yunchen Gong and Ms. Julia Copeland for sequencing data processing, quality control, and analysis.

## ETHICS APPROVAL AND CONSENT TO PARTICIPATE

The 2004 Pelotas Birth cohort study was approved by the Research Ethics Committee of the Faculty of Medicine at the Universidade Federal de Pelotas for all follow-ups and written informed consent was obtained from the parents (CEP 96.020-360, 96.030-001, 70.750-521). Approval for the analyses included in this work was granted by the Ethics Committee from the University of Toronto (REB #36176) and the Hospital for Sick Children, Toronto (REB #1000059180).

## REFERENCES

Agans, R., Rigsbee, L., Kenche, H., Michail, S., Khamis, H. J., & Paliy, O. (2011). Distal gut microbiota of adolescent children is different from that of adults. FEMS Microbiol Ecol, 77(2), 404–412. https://doi.org/10.1111/j.1574-6941.2011.01120.x

Anderson, M. J. (2001). A new method for non-parametric multivariate analysis of variance. Austral Ecology, 26(1), 32–46. https://doi.org/10.1111/j.1442-9993.2001.01070.pp.x

Balasubramaniam, C., Mallappa, R. H., Singh, D. K., Chaudhary, P., Bharti, B., Muniyappa, S. K., & Grover, S. (2021). Gut bacterial profile in Indian children of varying nutritional status: A comparative pilot study. European Journal of Nutrition, 60(7), 3971–3985. https://doi.org/10.1007/s00394-021-02571-7

Barros, A. J., da Silva dos Santos, I., Victora, C. G., Albernaz, E. P., Domingues, M. R., Timm, I. K., Matijasevich, A., Bertoldi, A. D., & Barros, F. C. (2006). The 2004 Pelotas birth cohort: Methods and description. Rev Saude Publica, 40(3), 402–413. https://doi.org/10.1590/s0034-89102006000300007

Bokulich, N. A., Chung, J., Battaglia, T., Henderson, N., Jay, M., Li, H., D. Lieber, A., Wu, F., Perez-Perez, G. I., Chen, Y., Schweizer, W., Zheng, X., Contreras, M., Dominguez-Bello, M. G., & Blaser, M. J. (2016). Antibiotics, birth mode, and diet shape microbiome maturation during early life. Science Translational Medicine, 8(343), 343ra82–343ra82. https://doi.org/10.1126/scitranslmed.aad7121

Bolyen, E., Rideout, J. R., Dillon, M. R., Bokulich, N. A., Abnet, C. C., Al-Ghalith, G. A., Alexander, H., Alm, E. J., Arumugam, M., Asnicar, F., Bai, Y., Bisanz, J. E., Bittinger, K., Brejnrod, A., Brislawn, C. J., Brown, C. T., Callahan, B. J., Caraballo-Rodríguez, A. M., Chase, J., … Caporaso, J. G. (2019). Reproducible, interactive, scalable and extensible microbiome data science using QIIME 2. Nature Biotechnology, 37(8), Article 8. https://doi.org/10.1038/s41587-019-0209-9

Calcaterra, V., Rossi, V., Massini, G., Regalbuto, C., Hruby, C., Panelli, S., Bandi, C., & Zuccotti, G. (2022). Precocious puberty and microbiota: The role of the sex hormone–gut microbiome axis. Frontiers in Endocrinology, 13. https://www.frontiersin.org/articles/10.3389/fendo.2022.1000919

Castaner, O., Goday, A., Park, Y.-M., Lee, S.-H., Magkos, F., Shiow, S.-A. T. E., & Schröder, H. (2018). The Gut Microbiome Profile in Obesity: A Systematic Review. International Journal of Endocrinology, 2018, 4095789. https://doi.org/10.1155/2018/4095789

Chao, A. (1984). Nonparametric Estimation of the Number of Classes in a Population. Scandinavian Journal of Statistics, 11(4), 265–270.

Edgar, R. C. (2010). Search and clustering orders of magnitude faster than BLAST. Bioinformatics, 26(19), 2460–2461. https://doi.org/10.1093/bioinformatics/btq461

Edgar, R. C. (2013). UPARSE: Highly accurate OTU sequences from microbial amplicon reads. Nature Methods, 10(10), Article 10. https://doi.org/10.1038/nmeth.2604

Edgar, R. C. (2016). *UNOISE2: Improved error-correction for Illumina 16S and ITS amplicon sequencing* [Preprint]. Bioinformatics. https://doi.org/10.1101/081257

Habicht, J. P. (1974). Estandarización de metodos epidemiológicos cuantitativos sobre el terreno. Boletín de La Oficina Sanitaria Panamericana (OSP);76(5),Mayo 1974. https://iris.paho.org/handle/10665.2/10766

Hollister, E. B., Riehle, K., Luna, R. A., Weidler, E. M., Rubio-Gonzales, M., Mistretta, T. A., Raza, S., Doddapaneni, H. V., Metcalf, G. A., Muzny, D. M., Gibbs, R. A., Petrosino, J. F., Shulman, R. J., & Versalovic, J. (2015). Structure and function of the healthy pre-adolescent pediatric gut microbiome. Microbiome, 3, 36. https://doi.org/10.1186/s40168-015-0101-x

Kim, Y. S., Unno, T., Kim, B.-Y., & Park, M.-S. (2020). Sex Differences in Gut Microbiota. The World Journal of Men’s Health, 38(1), 48–60. https://doi.org/10.5534/wjmh.190009

Korpela, K., Kallio, S., Salonen, A., Hero, M., Kukkonen, A. K., Miettinen, P. J., Savilahti, E., Kohva, E., Kariola, L., Suutela, M., Tarkkanen, A., de Vos, W. M., Raivio, T., & Kuitunen, M. (2021). Gut microbiota develop towards an adult profile in a sex-specific manner during puberty. Scientific Reports, 11(1), 23297. https://doi.org/10.1038/s41598-021-02375-z

Lopes Tdo, V., Cyrillo, D. C., Giuntini, E. B., Lajolo, F. M., & Menezes, E. W. (2015). [Brazilian Food Composition Database (TBCA-USP): Data compilation to serve the public good]. Arch Latinoam Nutr, 65(3), 186–192.

López-Domínguez, L., Bassani, D. G., Bourdon, C., Massara, P., Santos, I. S., Matijasevich, A., Barros, A. J. D., Comelli, E. M., & Bandsma, R. H. J. (2023). A novel shape-based approach to identify gestational age-adjusted growth patterns from birth to 11 years of age. Scientific Reports, 13(1), Article 1. https://doi.org/10.1038/s41598-023-28485-4

Lozupone, C. A., Hamady, M., Kelley, S. T., & Knight, R. (2007). Quantitative and Qualitative β Diversity Measures Lead to Different Insights into Factors That Structure Microbial Communities. Applied and Environmental Microbiology, 73(5), 1576–1585. https://doi.org/10.1128/AEM.01996-06

Lozupone, C., & Knight, R. (2005). UniFrac: A New Phylogenetic Method for Comparing Microbial Communities. Applied and Environmental Microbiology, 71(12), 8228–8235. https://doi.org/10.1128/AEM.71.12.8228-8235.2005

Mandal, S., Van Treuren, W., White, R. A., Eggesbø, M., Knight, R., & Peddada, S. D. (2015). Analysis of composition of microbiomes: A novel method for studying microbial composition. Microbial Ecology in Health and Disease, 26. https://doi.org/10.3402/mehd.v26.27663

Markle, J. G. M., Frank, D. N., Mortin-Toth, S., Robertson, C. E., Feazel, L. M., Rolle-Kampczyk, U., von Bergen, M., McCoy, K. D., Macpherson, A. J., & Danska, J. S. (2013). Sex differences in the gut microbiome drive hormone-dependent regulation of autoimmunity. Science (New York, N.Y.), 339(6123), 1084–1088. https://doi.org/10.1126/science.1233521

Marshall, W. A., & Tanner, J. M. (1969). Variations in pattern of pubertal changes in girls. Arch Dis Child, 44(235), 291–303.

Marshall, W. A., & Tanner, J. M. (1970). Variations in the pattern of pubertal changes in boys. Arch Dis Child, 45(239), 13–23.

Matsudo, S. M. M., & Matsudo, V. K. R. (1994). Self-assessment and physician assessment of sexual maturation in Brazilian boys and girls: Concordance and reproducibility. Am J Hum Biol, 6(4), 451–455. https://doi.org/10.1002/ajhb.1310060406

Méndez-Salazar, E. O., Ortiz-López, M. G., Granados-Silvestre, M. de los Á., Palacios-González, B., & Menjivar, M. (2018). Altered Gut Microbiota and Compositional Changes in Firmicutes and Proteobacteria in Mexican Undernourished and Obese Children. Frontiers in Microbiology, 9. https://doi.org/10.3389/fmicb.2018.02494

Norris, S. A., Frongillo, E. A., Black, M. M., Dong, Y., Fall, C., Lampl, M., Liese, A. D., Naguib, M., Prentice, A., Rochat, T., Stephensen, C. B., Tinago, C. B., Ward, K. A., Wrottesley, S. V., & Patton, G. C. (2022). Nutrition in adolescent growth and development. The Lancet, 399(10320), 172–184. https://doi.org/10.1016/S0140-6736(21)01590-7

Radjabzadeh, D., Boer, C. G., Beth, S. A., van der Wal, P., Kiefte-De Jong, J. C., Jansen, M. A. E., Konstantinov, S. R., Peppelenbosch, M. P., Hays, J. P., Jaddoe, V. W. V., Ikram, M. A., Rivadeneira, F., van Meurs, J. B. J., Uitterlinden, A. G., Medina-Gomez, C., Moll, H. A., & Kraaij, R. (2020). Diversity, compositional and functional differences between gut microbiota of children and adults. Scientific Reports, 10(1), 1040. https://doi.org/10.1038/s41598-020-57734-z

Rasmussen, A. R., Wohlfahrt-Veje, C., Tefre de Renzy-Martin, K., Hagen, C. P., Tinggaard, J., Mouritsen, A., Mieritz, M. G., & Main, K. M. (2015). Validity of self-assessment of pubertal maturation. Pediatrics, 135(1), 86–93. https://doi.org/10.1542/peds.2014-0793

Reyna, M. E., Petersen, C., Dai, D. L. Y., Dai, R., Becker, A. B., Azad, M. B., Miliku, K., Lefebvre, D. L., Moraes, T. J., Mandhane, P. J., Boutin, R. C. T., Finlay, B. B., Simons, E., Kozyrskyj, A. L., Lou, W., Turvey, S. E., & Subbarao, P. (2022). Longitudinal body mass index trajectories at preschool age: Children with rapid growth have differential composition of the gut microbiota in the first year of life. International Journal of Obesity, 46(7), Article 7. https://doi.org/10.1038/s41366-022-01117-z

Ringel-Kulka, T., Cheng, J., Ringel, Y., Salojarvi, J., Carroll, I., Palva, A., de Vos, W. M., & Satokari, R. (2013). Intestinal microbiota in healthy U.S. young children and adults—A high throughput microarray analysis. PLoS One, 8(5), e64315. https://doi.org/10.1371/journal.pone.0064315

Robertson, R. C., Manges, A. R., Finlay, B. B., & Prendergast, A. J. (2019). The Human Microbiome and Child Growth – First 1000 Days and Beyond. Trends in Microbiology, 27(2), 131–147. https://doi.org/10.1016/j.tim.2018.09.008

Rognes, T., Flouri, T., Nichols, B., Quince, C., & Mahé, F. (2016). VSEARCH: A versatile open source tool for metagenomics. PeerJ, 4, e2584. https://doi.org/10.7717/peerj.2584

Sanchez, G. (2013). *Plspm* [R]. https://github.com/gastonstat/plspm

Santos, I. S., Barros, A. J., Matijasevich, A., Domingues, M. R., Barros, F. C., & Victora, C. G. (2011). Cohort profile: The 2004 Pelotas (Brazil) birth cohort study. Int J Epidemiol, 40(6), 1461–1468. https://doi.org/10.1093/ije/dyq130

Santos, I. S., Barros, A. J., Matijasevich, A., Zanini, R., Chrestani Cesar, M. A., Camargo-Figuera, F. A., Oliveira, I. O., Barros, F. C., & Victora, C. G. (2014). Cohort profile update: 2004 Pelotas (Brazil) Birth Cohort Study. Body composition, mental health and genetic assessment at the 6 years follow-up. Int J Epidemiol, 43(5), 1437–1437a-f. https://doi.org/10.1093/ije/dyu144

Schmitz, K. E., Hovell, M. F., Nichols, J. F., Irvin, V. L., Keating, K., Simon, G. M., Gehrman, C., & Jones, K. L. (2004). A Validation Study of Early Adolescents’ Pubertal Self-Assessments. The Journal of Early Adolescence, 24(4), 357–384. https://doi.org/10.1177/0272431604268531

Schneider, B. C., Motta, J. V., Muniz, L. C., Bielemann, R. M., Madruga, S. W., Orlandi, S. P., Gigante, D. P., & Assuncao, M. C. (2016). Design of a digital and self-reported food frequency questionnaire to estimate food consumption in adolescents and young adults: Birth cohorts at Pelotas, Rio Grande do Sul, Brazil. Rev Bras Epidemiol, 19(2), 419–432. https://doi.org/10.1590/1980-5497201600020017

Shannon, C. E. (1948). A Mathematical Theory of Communication. Bell System Technical Journal, 27(3), 379–423. https://doi.org/10.1002/j.1538-7305.1948.tb01338.x

Shin, J.-H., Park, Y.-H., Sim, M., Kim, S.-A., Joung, H., & Shin, D.-M. (2019). Serum level of sex steroid hormone is associated with diversity and profiles of human gut microbiome. Research in Microbiology, 170(4–5), 192–201. https://doi.org/10.1016/j.resmic.2019.03.003

Silveira Schuch, H., Venâncio Fernandes Dantas, R., Menezes Seerig, L., S. Santos, I., Matijasevich, A., J.D. Barros, A., Glazer Peres, K., Peres, M. A., & Demarco, F. F. (2021). Socioeconomic inequalities explain the association between source of drinking water and dental caries in primary dentition. Journal of Dentistry, 106, 103584. https://doi.org/10.1016/j.jdent.2021.103584

Singh, R. K., Chang, H. W., Yan, D., Lee, K. M., Ucmak, D., Wong, K., Abrouk, M., Farahnik, B., Nakamura, M., Zhu, T. H., Bhutani, T., & Liao, W. (2017). Influence of diet on the gut microbiome and implications for human health. J Transl Med, 15(1), 73. https://doi.org/10.1186/s12967-017-1175-y

Stanislawski, M. A., Dabelea, D., Wagner, B. D., Iszatt, N., Dahl, C., Sontag, M. K., Knight, R., Lozupone, C. A., & Eggesbø, M. (2018). Gut Microbiota in the First 2 Years of Life and the Association with Body Mass Index at Age 12 in a Norwegian Birth Cohort. MBio, 9(5). https://doi.org/10.1128/mBio.01751-18

Stewart, C. J., Ajami, N. J., O’Brien, J. L., Hutchinson, D. S., Smith, D. P., Wong, M. C., Ross, M. C., Lloyd, R. E., Doddapaneni, H., Metcalf, G. A., Muzny, D., Gibbs, R. A., Vatanen, T., Huttenhower, C., Xavier, R. J., Rewers, M., Hagopian, W., Toppari, J., Ziegler, A.-G., … Petrosino, J. F. (2018). Temporal development of the gut microbiome in early childhood from the TEDDY study. Nature, 562(7728), 583–588. https://doi.org/10.1038/s41586-018-0617-x

Subramanian, S., Huq, S., Yatsunenko, T., Haque, R., Mahfuz, M., Alam, M. A., Benezra, A., DeStefano, J., Meier, M. F., Muegge, B. D., Barratt, M. J., VanArendonk, L. G., Zhang, Q., Province, M. A., Jr, W. A. P., Ahmed, T., & Gordon, J. I. (2014). Persistent gut microbiota immaturity in malnourished Bangladeshi children. Nature, 510(7505), 417–421. https://doi.org/10.1038/nature13421

Taylor, S. J., Whincup, P. H., Hindmarsh, P. C., Lampe, F., Odoki, K., & Cook, D. G. (2001). Performance of a new pubertal self-assessment questionnaire: A preliminary study. Paediatr Perinat Epidemiol, 15(1), 88–94.

Turnbaugh, P. J., Ridaura, V. K., Faith, J. J., Rey, F. E., Knight, R., & Gordon, J. I. (2009). The effect of diet on the human gut microbiome: A metagenomic analysis in humanized gnotobiotic mice. Sci Transl Med, 1(6), 6ra14. https://doi.org/10.1126/scitranslmed.3000322

U.S. Department of Agriculture, A. R. S., Nutrient Data Laboratory. (2017). USDA National Nutrient Database for Standard Reference, Release 28. Nutrient Data Laboratory Home Page. http://www.ars.usda.gov/nea/bhnrc/ndl

Valeri, F., & Endres, K. (2021). How biological sex of the host shapes its gut microbiota. Frontiers in Neuroendocrinology, 100912. https://doi.org/10.1016/j.yfrne.2021.100912

Vander Wyst, K. B., Ortega-Santos, C. P., Toffoli, S. N., Lahti, C. E., & Whisner, C. M. (2021). Diet, adiposity, and the gut microbiota from infancy to adolescence: A systematic review. Obesity Reviews: An Official Journal of the International Association for the Study of Obesity, 22(5), e13175. https://doi.org/10.1111/obr.13175

Vaz, J. S., Buffarini, R., Schneider, B. C., Bielemann, R. M., Gonçalves, H., & Assunção, M. C. F. (2021). Relative validity of a computer-based semi-quantitative FFQ for use in the Pelotas (Brazil) Birth Cohort Studies. Public Health Nutrition, 24(1), 34–42. https://doi.org/10.1017/S1368980020001196

WHO, W. H. O. (2010). Annex H, Sexual Maturity Rating (Tanner Staging) in Adolescents. In Antiretroviral Therapy for HIV Infection in Infants and Children: Towards Universal Access: Recommendations for a Public Health Approach: 2010 Revision (23741772).

Winpenny, E. M., van Sluijs, E. M. F., White, M., Klepp, K.-I., Wold, B., & Lien, N. (2018). Changes in diet through adolescence and early adulthood: Longitudinal trajectories and association with key life transitions. The International Journal of Behavioral Nutrition and Physical Activity, 15. https://doi.org/10.1186/s12966-018-0719-8

Wu, G. D., Chen, J., Hoffmann, C., Bittinger, K., Chen, Y. Y., & Keilbaugh, S. A. (2011). Linking long-term dietary patterns with gut microbial enterotypes. Science, 334. https://doi.org/10.1126/science.1208344

Yassour, M., Jason, E., Hogstrom, L. J., Arthur, T. D., Tripathi, S., Siljander, H., Selvenius, J., Oikarinen, S., Hyöty, H., Virtanen, S. M., Ilonen, J., Ferretti, P., Pasolli, E., Tett, A., Asnicar, F., Segata, N., Vlamakis, H., Lander, E. S., Huttenhower, C., … Xavier, R. J. (2018). Strain-Level Analysis of Mother-to-Child Bacterial Transmission during the First Few Months of Life. Cell Host & Microbe, 24(1), 146–154.e4. https://doi.org/10.1016/j.chom.2018.06.007

Yatsunenko, T., Rey, F. E., Manary, M. J., Trehan, I., Dominguez-Bello, M. G., Contreras, M., Magris, M., Hidalgo, G., Baldassano, R. N., Anokhin, A. P., Heath, A. C., Warner, B., Reeder, J., Kuczynski, J., Caporaso, J. G., Lozupone, C. A., Lauber, C., Clemente, J. C., Knights, D., … Gordon, J. I. (2012). Human gut microbiome viewed across age and geography. Nature, 486(7402), 222–227. https://doi.org/10.1038/nature11053

Yuan, X., Chen, R., Zhang, Y., Lin, X., & Yang, X. (2020). Gut microbiota: Effect of pubertal status. BMC Microbiology, 20(1), 334. https://doi.org/10.1186/s12866-020-02021-0

