## Supplemental figure and tables for "Childhood growth trajectory patterns are associated with the pubertal gut microbiota"

#### SUPPLEMENTARY INFORMATION

##### Supplementary table 1. Sub-selection of children from the 2004 Pelotas birth cohort

| **Classification** | **11 years** | | **Retention** | **13 years** | **Percentage of change** |
|---|---|---|---|---|---|
| | **Selected** | **Collected** | | **Collected** | |
| **Height-for-age z-score** | | | | | |
| *Stunting (< -2 z-score)* | 22 | 17 | 77% | 12 | 29% |
| *BMI-for-age z-score* | | | | | |
| **Severe wasting (< -3 z-score)** | 14 | 10 | 71% | 10 | 0% |
| *Wasting (< -2 z-score)* | 101 | 70 | 69% | 53 | 24% |
| *Normal weight* | 219 | 164 | 74% | 203 | 23% |
| *Overweight (< 2 z-score)* | 105 | 73 | 69% | 68 | 6% |
| *Obesity (> 3 z-score)* | 39 | 32 | 82% | 16 | 50% |
| **No information on weight/height** | | | | 3 | |
| **Total** | **500** | **366** | **76.4%** | **365** | |

**Supplementary table 2. List of food groups derived from Food Frequency Questionnaire**

| *Food group* | *Food items* |
|---|---|
| Beans | beans |
| Bread | whole wheat; white bread |
| Candies and sweets | candies; chocolate bar / bonbons; chocolate milk powder; fruit jam / jelly; dairy based sweets / pudding; ice cream |
| Coffee | coffee |
| Cookies and cake | sweet cookies; crackers; cake; cereal bars; breakfast cereals; granola |
| Milk and dairy products | cow's milk; yoghurt; cheese; soft cheese |
| Fast food and snacks | bauru / cheeseburger / hotdog; pizza; hamburger / nuggets; quibe; fried salty pastries; chips; canned vegetables (peas / corn / pickles); mayonnaise; sweet or savory popcorn |
| Fats | butter; margarine |
| Fish | fish |
| Fruits and fruit juices | avocado; pineapple; banana; guava; orange; apple / pear; papaya; mango; watermelon / cantaloupe; strawberry; peach; grape; artificial juice; natural juice |
| Meats and eggs | red meat; chicken; pork; gizzards / liver / heart / kidney; barbecue; eggs |
| Pastas and tubers | spaghetti; noodles; lasagna / ravioli / gnocchi; potato boiled / roasted / mashed; fried potato / polenta / cassava |
| Processed meats | bacon; sausage / frankfurter; mortadella / ham / salami |
| Rice | rice |
| Soda | sugar-sweetened and artificial-sweetened cola and other flavours |
| Sugar | sugar added to milk, juice, coffee, and tea |
| Teas | tea; chimarrao |
| Vegetable spices | garlic; onion; sweet pepper |
| Vegetables and legumes | pumpkin; lettuce; beet; carrot; cauliflower; chayote; cabbage; cucumber; cabbage; tomato; green beans; lentils / peas / chickpeas |

**Supplementary table 3. Correlation coefficients of alpha diversity of prenatal, sociodemographic and anthropometric variables**

| | *All* | | | | *Female* | | | | *Male* | | | |
|---|---|---|---|---|---|---|---|---|---|---|---|---|
| | *Chao1* | | *Shannon* | | *Chao1* | | *Shannon* | | *Chao1* | | *Shannon* | |
| | *Rho* | *p-val* | *Rho* | *p-val* | *Rho* | *p-val* | *Rho* | *p-val* | *Rho* | *p-val* | *Rho* | *p-val* |
| **Age (years)** | 0.18 | **0.001** | 0.06 | 0.28 | 0.12 | 0.14 | 0.01 | 0.88 | 0.23 | **0.001** | 0.11 | 0.15 |
| **Maternal age (years)** | -0.13 | **0.01** | -0.12 | **0.03** | -0.23 | **0.004** | -0.24 | **0.002** | -0.03 | 0.70 | 0.01 | 0.93 |
| **Maternal education (years)** | -0.18 | **0.0008** | -0.15 | **0.005** | -0.11 | 0.20 | -0.05 | 0.53 | -0.27 | **0.0003** | -0.24 | **0.001** |
| **Maternal BMI (kg/m2)** | -0.06 | 0.34 | 0.02 | 0.77 | -0.05 | 0.62 | -0.05 | 0.63 | -0.08 | 0.38 | 0.09 | 0.33 |
| **Dietary patterns** | | | | | | | | | | | | |
| *Processed* | 0.10 | 0.06 | 0.07 | 0.19 | 0.02 | 0.81 | 0.02 | 0.82 | 0.16 | 0.04 | 0.11 | 0.13 |
| *Coffee, Tea, Vegetables* | 0.19 | **0.0005** | 0.07 | 0.22 | 0.16 | 0.05 | 0.07 | 0.39 | 0.23 | **0.002** | 0.06 | 0.44 |
| *Common-Brazilian* | 0.15 | **0.005** | 0.14 | **0.01** | 0.20 | **0.01** | 0.20 | **0.01** | 0.10 | 0.20 | 0.08 | 0.29 |
| **Age of introduction of solids (months)** | 0.05 | 0.38 | 0.03 | 0.65 | 0.06 | 0.50 | 0.05 | 0.57 | 0.04 | 0.57 | 0.03 | 0.74 |
| **BMI-for-age z-score** | -0.21 | **0.0001** | -0.16 | **0.005** | -0.03 | 0.69 | -0.11 | 0.18 | -0.36 | **0.00** | 0.84 | **0.01** |
| **Height-for-age z-score** | -0.05 | 0.35 | -0.03 | 0.60 | -0.01 | 0.91 | -0.01 | 0.92 | -0.10 | 0.17 | -0.02 | 0.84 |
| **Fat mass at 11 years (%)** | -0.31 | **0.00** | -0.26 | **0.00** | -0.27 | **0.0009** | -0.26 | **0.001** | -0.37 | **0.00** | -0.24 | **0.002** |

Spearman correlations

**Supplementary table 4. Comparison of gut microbiota beta-diversity and variables of prenatal, sociodemographic and anthropometric variables**

| | Unweighted UNIFRAC | | | Weighted UNIFRAC | | |
|---|---|---|---|---|---|---|
| | All (n=331) | Females (n=154) | Males (n=77) | All (n=331) | Females (n=154) | Males (n=77) |
| **Skin colour** | | | | | | |
| *Black vs brown* | 0.79 | 0.27 | 0.78 | 0.82 | 0.93 | 0.61 |
| *Black vs other* | 0.13 | 0.28 | 0.05 | 0.11 | 0.93 | 0.14 |
| *Black vs white* | **0.006** | **0.01** | **0.01** | **0.02** | 0.05 | 0.14 |
| *Brown vs other* | 0.27 | 0.56 | 0.12 | 0.20 | 0.93 | 0.14 |
| *Brown vs white* | **0.006** | 0.17 | **0.006** | **0.02** | 0.05 | 0.14 |
| *Other vs white* | **0.02** | 0.41 | **0.009** | 0.33 | 0.93 | 0.14 |
| **Income** | | | | | | |
| *Quintile 1 vs 2* | 0.60 | 0.38 | 0.81 | 0.29 | 0.13 | 0.78 |
| *Quintile 1 vs 3* | 0.34 | 0.88 | 0.72 | **0.03** | 0.16 | 0.08 |
| *Quintile 1 vs 4* | **0.01** | 0.38 | 0.05 | 0.07 | 0.76 | 0.06 |
| *Quintile 1 vs 5* | **0.003** | **0.01** | **0.01** | **0.01** | 0.09 | **0.04** |
| *Quintile 2 vs 3* | 0.23 | 0.41 | 0.37 | 0.29 | 0.76 | 0.06 |
| *Quintile 2 vs 4* | 0.10 | 0.38 | **0.02** | 0.44 | 0.25 | 0.05 |
| *Quintile 2 vs 5* | **0.003** | **0.03** | **0.01** | 0.03 | 0.25 | **0.02** |
| *Quintile 3 vs 4* | 0.07 | 0.38 | 0.36 | 0.29 | 0.33 | 0.59 |
| *Quintile 3 vs 5* | **0.003** | **0.01** | 0.05 | 0.13 | 0.13 | 0.59 |
| *Quintile 4 vs 5* | **0.01** | 0.06 | 0.35 | 0.11 | 0.13 | 0.59 |
| **Maternal smoking** | | | | | | |
| *No vs yes* | **0.004** | 0.08 | **0.009** | 0.12 | 0.43 | 0.23 |
| **Type of birth** | | | | | | |
| *Vaginal vs caesarean* | **0.008** | 0.21 | **0.002** | 0.23 | 0.54 | 0.27 |
| **Breastfeeding at 3 months** | | | | | | |
| *Exclusive vs no* | 0.58 | 0.94 | 0.68 | 0.90 | 0.75 | 0.73 |
| *Exclusive vs partial* | 0.42 | 0.91 | 0.75 | 0.44 | 0.78 | 0.73 |
| *Exclusive vs predominant* | 0.58 | 0.91 | 0.89 | 0.87 | 0.78 | 0.73 |
| *No vs partial* | 0.58 | 0.91 | 0.89 | 0.44 | 0.47 | 0.83 |
| *No vs predominant* | 0.56 | 0.91 | 0.68 | 0.87 | 0.75 | 0.83 |
| *Predominant vs partial* | 0.42 | 0.91 | 0.68 | 0.71 | 0.75 | 0.73 |
| **Breastfeeding at 12 months** | | | | | | |
| *No vs yes* | 0.44 | 0.46 | 0.37 | 0.65 | 0.16 | 0.88 |
| **BMI-for-age z-score** | | | | | | |
| *Normal vs overweight* | **0.006** | 0.40 | **0.003** | 0.12 | 0.20 | 0.22 |

| | | | | | | |
|---|---|---|---|---|---|---|
| *Normal vs wasting* | 0.75 | 0.76 | 0.35 | 0.20 | 0.98 | 0.22 |
| *Overweight vs wasting* | **0.04** | 0.49 | **0.02** | 0.14 | 0.98 | 0.22 |
| **Height-for-age z-score** | | | | | | |
| *Stunting vs normal* | 0.05 | 0.91 | 0.14 | 0.06 | 0.38 | 0.14 |

### Supplementary figure 1. Alpha diversity of prenatal, sociodemographic and anthropometric variables

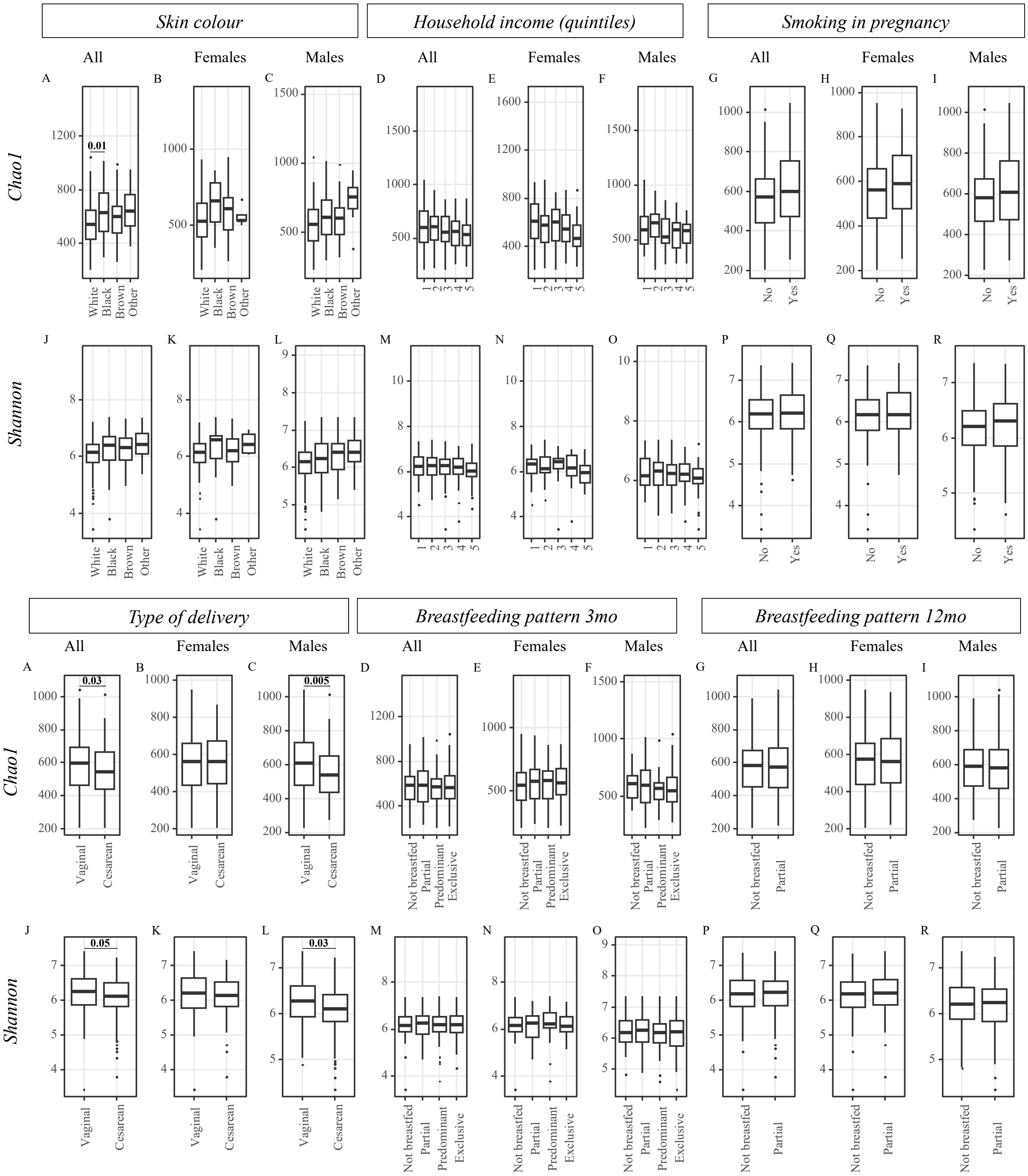

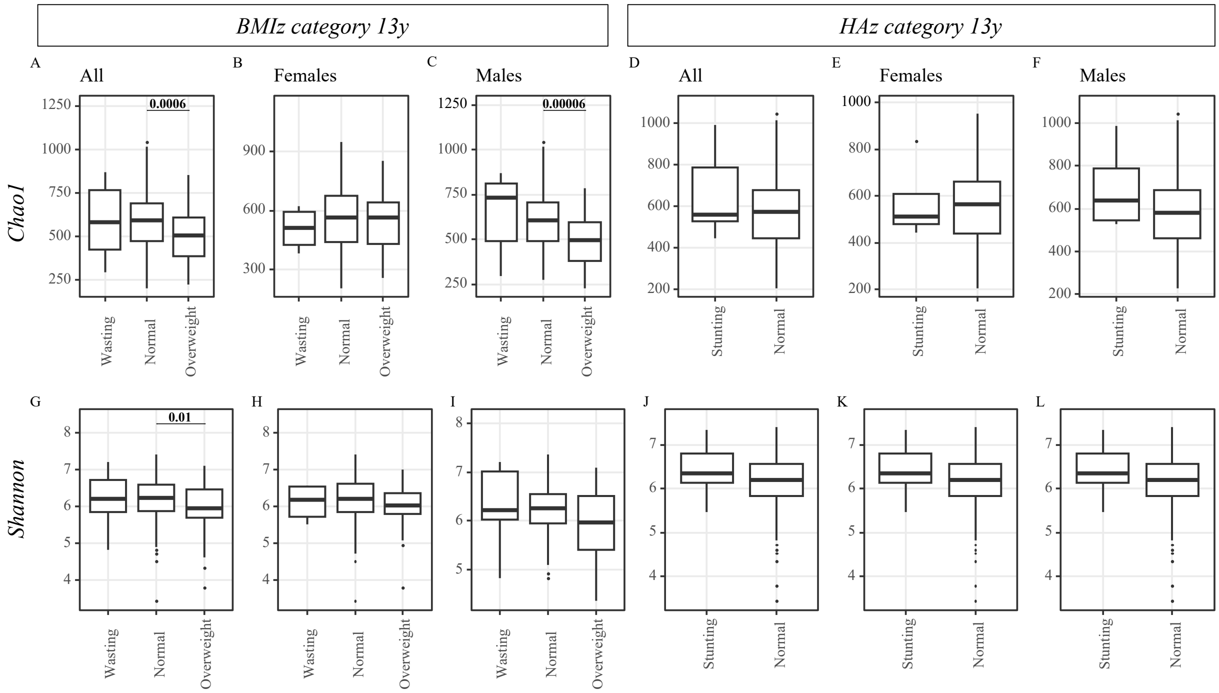
BMIz category 13y
HAz category 13y
A
All
B
Females
C
Males
D
All
E
Females
F
Males
Chao1
Shannon
0.0006
0.00006
0.01
G
H
I
J
K
L
Wasting
Normal
Overweight
Stunting
Normal
